# Polygenic Risk Scores for Alzheimer’s Disease and Mild Cognitive Impairment in Hispanics/Latinos in the U.S: The Study of Latinos – Investigation of Neurocognitive Aging

**DOI:** 10.1101/2021.01.08.21249413

**Authors:** Tamar Sofer, Nuzulul Kurniansyah, Einat Granot-Hershkovitz, Matthew O. Goodman, Wassim Tarraf, Iris Broce, Richard B. Lipton, Martha Daviglus, Melissa Lamar, Sylvia Wassertheil-Smoller, Jianwen Cai, Charles S. DeCarli, Hector M. Gonzalez, Myriam Fornage

## Abstract

**Introduction:** Polygenic Risk Score (PRS) are powerful summaries of genetic risk alleles that can potentially be used to predict disease outcomes and guide treatment decisions. Hispanics/Latinos suffer from higher rates of Alzheimer’s Disease (AD) and Mild Cognitive Impairment (MCI) compared to non-Hispanic Whites, yet the strongest known genetic risk factor for AD, *APOE*-*ϵ*4 allele, has weak association with AD in Hispanics/Latinos. We evaluated PRS constructed based on Genome-Wide Association Studies (GWAS) of AD in predicting MCI in Hispanics/Latinos when accounting for *APOE* alleles and variants.

**Methods:** We used summary statistics from four GWAS of AD to construct PRS that predict MCI in 4,189 diverse Hispanics/Latinos (mean age 63 years, 47% males) from the Study of Latinos-Investigation of Neurocognitive Aging. We assessed the PRS associations with MCI in the combined set of people and in groups defined by genetic ancestry and Hispanic/Latino background, and when including and excluding single nucleotide polymorphisms (SNPs) from the *APOE* gene region.

**Results:** A PRS constructed based on GWAS of AD in the FINNGEN Biobank was associated with MCI (OR = 1.34, 95% CI [1.15, 1.55]), and its association was mostly driven by 158 *APOE* region SNPs. A PRS constructed based on a multi-ethnic AD GWAS was associated with MCI (OR=1.22, 95% CI [1.08, 1.37]) without including any *APOE* region SNPs. *APOE*-*ϵ*4 and *APOE*-*ϵ*2 alleles were not associated with MCI.

**Discussion:** A combination of *APOE* region SNPs is associated with MCI in Hispanics/Latinos despite *APOE*-*ϵ*4 and *APOE*-*ϵ*2 alleles not being associated with MCI.

## Introduction

Hispanics/Latinos are the largest growing minority in the U.S., projected to represent 28.6% of the U.S. population by 2060 (Colby & Ortman 2015). Rates of Alzheimer’s disease and related dementia (ADRD) and mild cognitive impairment (MCI), which often precedes ADRD, are higher in Hispanics/Latinos compared to European Americans (Choi et al. 2018a, Mayeda et al. 2016, Weden et al. 2017). However, the strongest known genetic risk factors for ADRD, the *APOE*-*ϵ*4 allele (Genin et al. 2011), has weaker association in Hispanics/Latinos compared to individuals of European ancestry (Farrer 1997), and was not associated with MCI in recent studies from the Study of Latinos – Investigation of Cognitive Aging (SOL-INCA) (González et al. 2019b, Granot-Hershkovitz et al. 2020). Polygenic Risk Score (PRS) are aggregated summaries of genetic data, generally defined as weighted sums of counts of risk alleles for a particular health outcome across the genome. Thus, by collecting information genome-wide, they may assist in explaining the genetic association of ADRD and MCI beyond the *APOE* alleles and perhaps help elucidate ADRD disparities to some extent. Further, as PRS are more developed, they are starting to become useful for risk prediction (Lambert et al. 2019), potentially leading to disease prevention (Chatterjee et al. 2016), e.g. by risk stratification, and by personalizing interventions (Torkamani et al. 2018). Thus, applying PRS to evaluate personalized susceptibility for ADRD and MCI may be a useful target that will ultimately improve these outcomes among Hispanics/Latinos.

PRS are typically constructed based on summary statistics from genome-wide association studies (GWAS). It is already known that, to be useful for PRS construction, a GWAS need to have a large enough sample size (Choi et al. 2018b). By now, published GWAS of AD are available, for example, from the International Genomics of Alzheimer’s Project (IGAP; (Kunkle et al. 2019, Lambert et al. 2013)), which performed GWAS in individuals of European ancestry, and from the Alzheimer’s Disease Genetics Consortium, which performed a trans-ethnic GWAS (Jun et al. 2017). Hispanics/Latinos are admixed, with European, African, and Amerindian ancestries, with varying degrees of admixture across groups defined by Hispanic/Latino background (Conomos et al. 2016). While no large GWAS matches the genetic ancestry composition of the SOL-INCA Hispanics/Latinos exactly, in the last 4 years of GWAS in the Hispanic Community Health Study/Study of Latinos (HCHS/SOL), we showed that many genetic loci for cardiometabolic and other complex traits identified in GWAS of other genetic ancestries also show associations in Hispanics/Latinos (Graff et al. 2017, Qi et al. 2017, Sofer et al. 2017b,a). PRS, however, do not generalize as well. For example, Grinde et al. (2019) studied generalization of PRS for anthropometric, blood pressure, and blood count phenotypes, constructed primary based on European ancestry GWAS, and showed that by constructing PRS using GWAS of both European ancestry and another GWAS in Hispanics/Latinos, the potential prediction capability of the PRS substantially increases for blood pressure phenotypes, and most of the other phenotypes as well. Thus, we currently do not know how well PRS constructed based on non-Latino GWAS of AD will predict AD or MCI in Hispanics/Latinos.

The SOL-INCA is an ancillary study to the HCHS/SOL (González et al. 2019a), designed to study the development of AD and related dementia (ADRD) in U.S. Hispanics/Latinos. The average age at the SOL-INCA exam was 62, allowing for assessment of MCI, but not yet of ADRD, with MCI being defined using the National Institute on Aging– Alzheimer’s Association criteria for MCI syndromes (Albert et al. 2011). While we do not know yet how MCI will predict future ADRD, it is important to study how known genetic factors underlying ADRD, specifically, AD, may predict MCI in this populations. Recently, Logue et al. (2019) reported an association of a PRS constructed based on GWAS of AD from IGAP reported by Lambert et al. (2013) with MCI in a sample of middle aged (mean age 56) European Americans. Thus, there, both the discovery GWAS and the population with MCI were of the same genetic ancestry. It remains to study whether AD PRS, constructed based on GWAS in European ancestry individuals or in a multi-ethnic analysis, is associated with MCI in Hispanics/Latinos and whether *APOE* genotype impacts this association.

Here, we use summary statistics from four GWAS of Alzheimer’s disease and use them to develop PRS. Based on each GWAS, we select PRS that have the strongest potential to predict MCI in diverse U.S. Hispanics/Latinos from SOL-INCA. We estimate the associations of these PRS with MCI, examine whether the associations depend on genetic ancestry and the *APOE* genotype, by including and excluding *APOE* gene-region variants from the PRS, and by stratifying by APOE-*ϵ*4 carrier status. We also investigate the association of the PRS with change in global cognitive function and in specific domains. The conceptual organization of this study is described in Figure 1.

**Figure 1:**
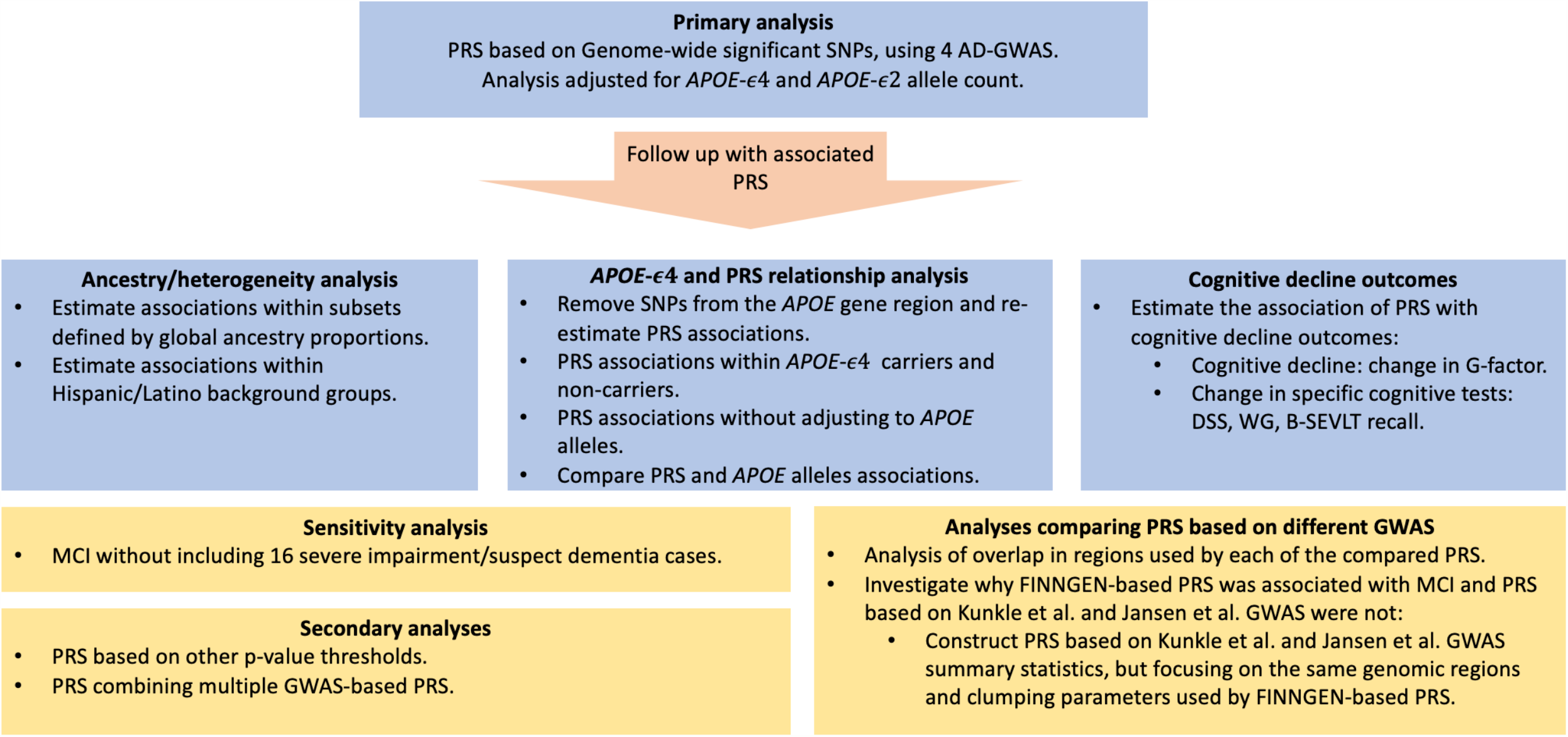
Analysis flowchart.

## Methods

### The Hispanic Community Health Study/Study of Latinos

The HCHS/SOL is a population-based longitudinal cohort study established to study risk and protective factors in cardiovascular disease development among U.S. Hispanics/Latinos (Sorlie et al. 2010). The study follows Hispanic/Latino participants from four metropolitan areas: Bronx NY, Miami FL, Chicago IL, and San Diego CA, with 16,415 participants aged 18-74 years examined in the baseline visit. Individuals were recruited via a probability sampling scheme from pre-defined census block units, chosen to provide diversity with respect to socioeconomic status as well as national origin or background (Lavange et al. 2010). Participants self-identified with six Hispanic/Latino background groups: Central American, South American, Mexican (Mainland groups, have high Amerindian genetic ancestry and low African ancestry), Cuban, Dominican, and Puerto-Rican (Caribbean group, have low Amerindian ancestry, and high African ancestry). At baseline, participants who were at least 45 years old and did not refuse nor had health limitations (n=9,714) were administered cognitive tests, including a Brief Spanish English verbal learning tests (B-SEVLT; (González et al. 2001)), assessing episodic learning and memory; word frequency test (also called word fluency, WF; (Lezak et al. 2012)), assessing verbal function; and digit symbol substitution test (DSS; (Wechsler 1981)), assessing processing speed and executive function. A second clinic visit occurred in 2014-2017, and during or after this visit, 6,377 participants who were eligible (completed neurocognitive testing during visit 1 and were at least 50 years old at visit 2) participated in the Study of Latinos-Investigation of Neurocognitive Aging (SOL-INCA), an ancillary study to the HCHS/SOL. SOL-INCA exams occurred, on average, 7 years after the baseline visit. Detailed information about the SOL-INCA exam and cognitive phenotyping is available in and González et al. (2019). In brief, the same cognitive battery from the baseline visit was applied and complemented with additional neurocognitive tests and instrumental activities of daily living questionnaire to assess functional status. In this study, we included n=4,256 individuals who participated in both the SOL-INCA study and were genotyped (as detailed below). All individuals provided written informed consent at their recruitment site.

### Cognitive phenotypes

Our primary phenotype was MCI, defined according to the National Institute on Aging-Alzheimer’s Association (NIA-AA) criteria (Albert et al. 2011), as detailed in González et al. (2019). In brief, a participant was classified with MCI if the three conditions were satisfied: (a) any cognitive test score in the range −1 to −2 SD (standard deviation) of the SOL-INCA robust norms adjusted for age, sex, education, and Picture Vocabulary Test scores, (b) a global measure of cognition declined by more than 0.055 SD per year between the visits, and (c) participant self-reported cognitive decline based on an Everyday Cognition questionnaire. Finally, we also included in the MCI group 16 individuals with severe impairment/suspect dementia, defined by a “cognitive deficit” of any cognitive test score lower than −2 SD compared to the internal norms, and more than minimal functional impairment according to the instrumental daily living questionnaire.

In secondary analysis, we also report PRS associations with changes in specific test scores measured in both the baseline and the SOL-INCA visits: B-SEVLT, WF, and DSS, and with a cognitive decline variable computed as a change in G-factor. To compute this variable, we used the baseline cognitive tests (B-SEVLT recall, DSS, and WF) restricted to the set of people who participated in the SOL-INCA exam. We computed the mean and standard deviation (SD) of each of the tests during the baseline visit while accounting for study design and sampling into the SOL-INCA study. We used these estimated means and SDs to scale the cognitive test values in each of the baseline and SOL-INCA exams (we subtracted the scores of each test by the corresponding mean and divided by the corresponding SD). We then computed principal components based on the standardized baseline exam test scores and obtained loadings for each of the three (standardized) tests. We used the loadings to compute the first principel component scores in both the baseline and the SOL-INCA visit to obtain G_baseline_ and G_inca_. The change in global cognitive function was computed as G_change_ = G_inca_ - G_baseline_. Negative values indicate decline in global cognitive function between the baseline and the SOL-INCA exams. Note that some values were positive, but the trend across all participants was of decline.

### Genotyping and imputation in HCHS/SOL

Blood was drawn from HCHS/SOL participants during the baseline exam. Individuals who consented to genetic studies were genotyped using an Illumina Omni2.5M array, which included 150,000 custom-selected Single Nucleotide Polymorphisms (SNPs), including ancestry-informative and Amerindian-specific variants. Global ancestry proportions measuring the proportion of the genome inherited from European, African, and Amerindian ancestors, and genetic principal components, were computed as previously reported (Conomos et al. 2016). The genotypes were imputed to the Trans-Omics in Precision Medicine (TOPMed) freeze 5b reference panel as described in Kowalski et al. (2019). APOE genotyping was performed separately using a TaqMan assay as previously described (González et al. 2018).

### Discovery GWAS of Alzheimer’s Disease and related traits

We used summary statistics from four publicly available GWAS of AD to construct PRS, as reported in Table 1. We used the most recent IGAP GWAS (Kunkle et al. 2019), a meta-analysis of AD GWAS and GWAS of parental AD in the UK Biobank (Jansen et al. 2019), a GWAS of AD performed in the FINNGEN Biobank from June 16 2020 data release (name: G6_ALZHEIMER, long name: Alzheimer disease, tags: #G6,#GASTRO_CM; google cloud file name: finngen_r3_G6_ALZHEIMER.gz), and a multi-ethnic GWAS of AD (Jun et al. 2017). For the latter one, we did not have complete summary statistics, and we used a list of 596 SNPs that Jun et al. (2017) provided in their supplement. This list did not include SNPs from the *APOE* gene region.

**Table 1:** External GWAS used for AD PRS construction.

| GWAS name | Sample size (cases/controls) | Outcome | Race/ethnicity | Reference | Data download link |
| --- | --- | --- | --- | --- | --- |
| Kunkle et al.<br>(Kunkle et al. 2019) | 21,982 cases<br>41,944 controls (from stage 1) | AD | European ancestry | PMID:<br>30820047 | https://www.niagads.org/datasets/ng00075 |
| Jansen et al.<br>(Jansen et al. 2019) | 24,087 cases, 55,058 controls,<br>+ parental AD from UK<br>Biobank (47,793 proxy cases,<br>328,320 controls) | LOAD +<br>parental<br>AD | European ancestry | PMID:<br>30617256 | https://ctg.cncr.nl/software/summary_statistics |
| FINNGEN | 2,568 cases, 133,070 controls | AD | Finnish European |  | https://www.FINNGEN.fi/en/access_results<br>We used GWAS version published on June 16, 2020. |
| Jun et al. (Jun et al. 2017) | 15,579 cases, 17,690 controls | AD | Multi-ethnic: European ancestry, African American, Japanese, and Israeli Arab. | PMID:<br>28183528 | https://ars.els-cdn.com/content/image/1-s2.0-S1552526017300031-mmc3.xlsx |

### PRS construction

In primary analysis, we constructed PRS using genome-wide significant SNPs (p-value<5×10^−8^) in their respective GWAS (see Figure 1). We constructed PRS using the clump-and-threshold method implemented in the PRSice 2 software (Choi & O’Reilly 2019) with HCHS/SOL as the reference panel, and also without clumping, because linkage disequilibrium patterns differ between populations, so clumping may not be effective. We first lifted over summary statistics from genome build 37 to genome build 38, and removed summary statistics corresponding to SNPs with minor allele frequency lower than 1%. For PRS that use clumped SNPs, we applied PRSice to create PRS using clumping parameters R^2^ ∈ {0.1, 0.2, 0.3} and distances of 250Kb, 500Kb, and 100Kb. For a given set of R^2^ and distance clumping parameters, this means that once a SNP is selected, all other SNPs within that distance and with correlation higher than the set R^2^ are removed from consideration. In a secondary analysis, we also considered the following p-value thresholds on the summary statistics: {5 × 10^−8^, 10^−7^, 10^−6^, 10^−5^, 10^−4^, 10^−3^, 10^−2^, 0.1, 0.2, 0.3, 0.4, 0.5}.

### PRS selection

For a given p-value threshold, there are multiple PRS constructed, corresponding to the various clumping parameters. To select a single PRS from each GWAS, we removed related individuals from the data, followed by splitting it to 5 equally-sized subsets. We performed association analysis of each PRS with MCI and recorded the effect size. For each GWAS, we then chose the PRS that minimized the Coefficient of Variation (CV) across the 5 subsets. In secondary analysis considering a range of p-value thresholds to select SNPs to the PRS, for each GWAS we apply the same clumping parameters selected in the primary analysis, and report association testing results across PRS constructed according to the range of p-value thresholds.

### Primary association analysis

In primary analysis, we used the combined SOL-INCA population. PRS were standardized in association testing so that they had mean zero and variance 1, and estimated effect sizes are per 1 SD of the PRS. Standardizations was performed on the combined SOL-INCA population and were not performed again when considering subgroups. The association analyses used survey logistic (for MCI) and linear (for cognitive decline phenotypes) regression using the ‘survey’ R package (Lumley 2011), accounting for stratification and probability sampling, and further adjusted to sex, age at the baseline cognitive exam, time between the baseline exam and the SOL-INCA exam, education level (3-category variable: less than higher school diploma or GED, high school diploma or GED, or higher), study center, 5 principal components of genetic data, and for APOE-*ϵ*4 and APOE-*ϵ*2 allele counts. The sampling weights used in analyses were based on the SOL-INCA ancillary study, so that association analyses and sample characteristics represent the SOL-INCA target population. In a sensitivity analysis, we removed 16 individuals with severe cognitive deficit/suspect dementia and re-evaluated the PRS associations. Focusing on the PRS that were associated with MCI, we also performed additional analysis as described henceforth.

### Ancestry/heterogeneity analysis

We also report the association (effect sizes and p-values) of the MCI-associated PRS across subsets of individuals defined by proportion ancestry: at least 20% European, African, or Amerindian global ancestry, and associations within Hispanic/Latino background groups. When estimating associations within Hispanic/Latino background groups, we tested for heterogeneity using the Cochran Q test that accounts for covariance between effect estimates across the background group developed in Sofer et al. (2016), because individuals are potentially genetically related and potentially live in the same household despite having different Hispanic/Latino background group. To obtain estimates of the covariances between the group-specific effect estimates we fit a single logistic regression model that estimated the effect of the PRS in all background group jointly.

### Association with cognitive decline phenotypes

We estimated the association of the PRS identified in the primary analysis with cognitive decline phenotypes: change in specific test scores and change in G-factor (global cognitive decline).

### *APOE* and PRS relationship analysis

For the three GWAS with summary statistics containing SNPs in the *APOE* gene region, we removed all SNPs from this region as follows. We took the position of rs429358 (in genome build 38), and removed all SNPs within a region 1Mb region centered at rs429358. For each GWAS, we then constructed PRS again without these SNPs and using the same weights otherwise, and estimated the new PRS association. In another analysis, we used the PRS from the primary analysis (potentially including *APOE* region SNPs) and evaluated their association with MCI within carriers and non-carriers of an *APOE*-*ϵ*4 allele.

### PRS based on multiple GWAS

In a secondary analysis, we took two PRS that were associated with MCI and used them to construct a single PRS. We fitted the logistic regression model *logit* [Pr(*MCI* = 1)] = *β*_0_ + ***x***^*T*^ ***β***_*x*_ + *PRS*_1_*β*_1_+ *PRS*_2_ *β*_2_, where *β*_0_ is the intercept, ***x*** is a vector of adjusting variables (age, sex, etc.) and *β*_*x*_ their coefficient vector, and *β*_1,_ *β*_2_ are coefficient of the (standardized) *PRS*_1_, *PRS*_2,_ respectively. We obtained the estimates 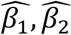, and used them to construct a new 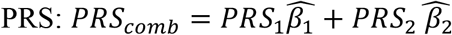. Because this PRS is overfitted to SOL-INCA (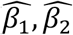, are optimized based on this dataset), this is a secondary analysis.

### Investigation of differences in MCI associations of different PRS

We performed two secondary analyses to investigate why some PRS were associated with MCI in Hispanics/Latinos and others were not. First, we studied the overlap between genomic regions represented by each of the PRS. A genomic region was identified according to a 1Mbp region around an index SNP. We identified genomic regions for the set of SNPs contributing to each PRS, and then counted overlapping and non-overlapping regions. Second, when one PRS based on GWAS individuals of European ancestry was associated with MCI, we constructed similar PRS based on other GWAS of European ancestry, i.e., by using the same clumping parameters and genomic regions to generate PRS, and used them in association analyses.

## Results

Table 2 characterizes the target population by Hispanic/Latino background group. At the SOL-INCA exam, the average age ranged from 62-65 years in the target population across Hispanic/Latino background groups, with the Cuban group being oldest on average with mean age 65.2. In many other characteristics, the background groups were quite heterogeneous: the Cuban group had the highest proportion of males (52.2%) while the Dominican group had the lowest (40.2%). The South American group were the most educated with 61.5% of the individuals having more than a high school diploma. Rates of MCI varied from 8% (South American) to 12.8% (Puerto Rican), and cognitive decline (G-factor change) ranged from 0.21 reduction (Mexican) to 0.07 reduction (South American). Global proportions of European ancestry ranged from 0.80 on average (Cuban) to 0.46 on average (Central American and Mexican).

**Table 2:**
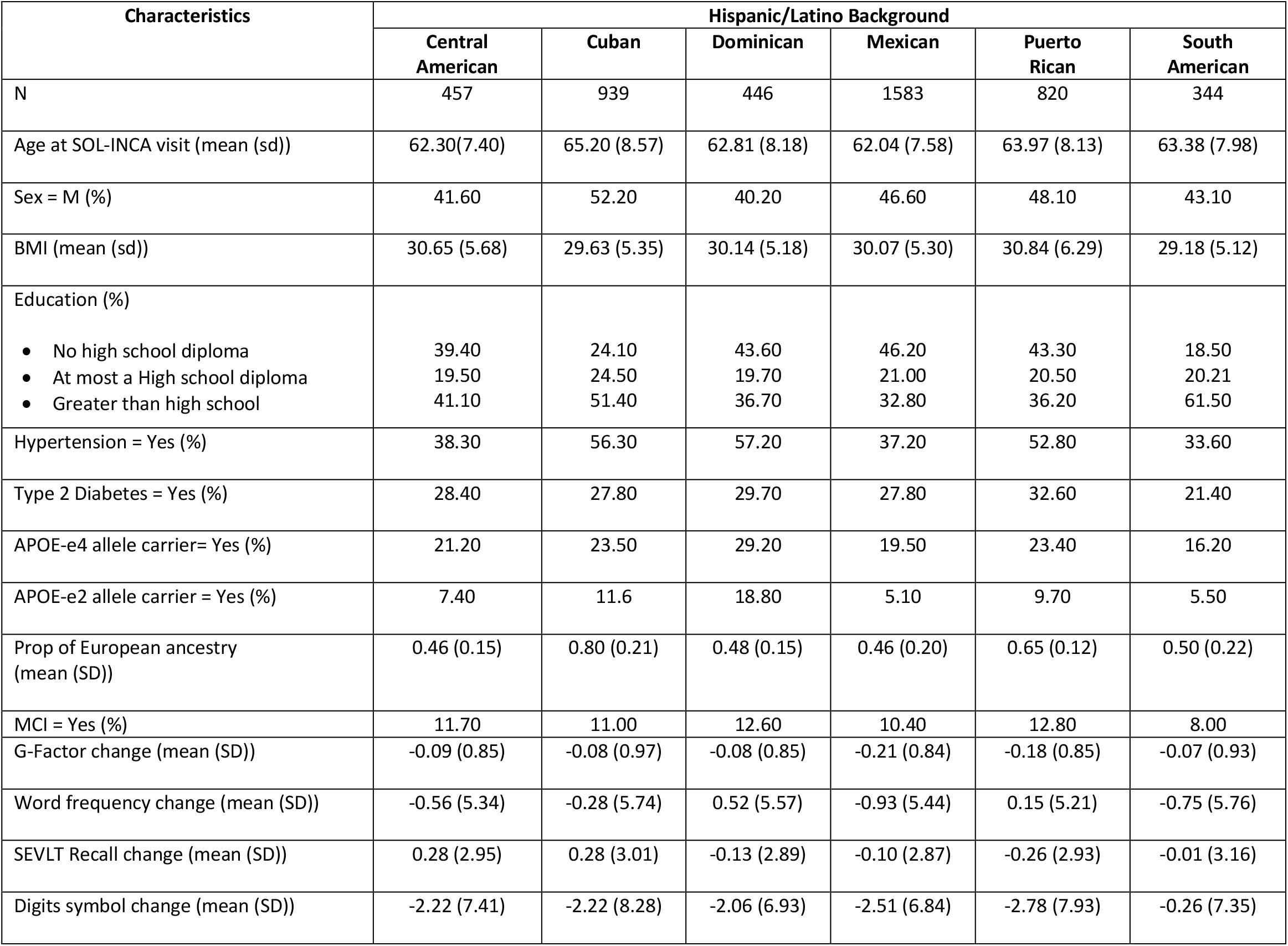
Participant characteristics by Hispanic/Latino background group (survey weighted).

| Characteristics | Hispanic/Latino Background |  |  |  |  |  |
| --- | --- | --- | --- | --- | --- | --- |
|  | Central American | Cuban | Dominican | Mexican | Puerto Rican | South American |
| N | 457 | 939 | 446 | 1583 | 820 | 344 |
| Age at SOL-INCA visit (mean (sd)) | 62.30(7.40) | 65.20 (8.57) | 62.81 (8.18) | 62.04 (7.58) | 63.97 (8.13) | 63.38 (7.98) |
| Sex = M (%) | 41.60 | 52.20 | 40.20 | 46.60 | 48.10 | 43.10 |
| BMI (mean (sd)) | 30.65 (5.68) | 29.63 (5.35) | 30.14 (5.18) | 30.07 (5.30) | 30.84 (6.29) | 29.18 (5.12) |
| Education (%) |  |  |  |  |  |  |
| • No high school diploma | 39.40 | 24.10 | 43.60 | 46.20 | 43.30 | 18.50 |
| • At most a High school diploma | 19.50 | 24.50 | 19.70 | 21.00 | 20.50 | 20.21 |
| • Greater than high school | 41.10 | 51.40 | 36.70 | 32.80 | 36.20 | 61.50 |
| Hypertension = Yes (%) | 38.30 | 56.30 | 57.20 | 37.20 | 52.80 | 33.60 |
| Type 2 Diabetes = Yes (%) | 28.40 | 27.80 | 29.70 | 27.80 | 32.60 | 21.40 |
| APOE-e4 allele carrier= Yes (%) | 21.20 | 23.50 | 29.20 | 19.50 | 23.40 | 16.20 |
| APOE-e2 allele carrier = Yes (%) | 7.40 | 11.6 | 18.80 | 5.10 | 9.70 | 5.50 |
| Prop of European ancestry (mean (SD)) | 0.46 (0.15) | 0.80 (0.21) | 0.48 (0.15) | 0.46 (0.20) | 0.65 (0.12) | 0.50 (0.22) |
| MCI = Yes (%) | 11.70 | 11.00 | 12.60 | 10.40 | 12.80 | 8.00 |
| G-Factor change (mean (SD)) | -0.09 (0.85) | -0.08 (0.97) | -0.08 (0.85) | -0.21 (0.84) | -0.18 (0.85) | -0.07 (0.93) |
| Word frequency change (mean (SD)) | -0.56 (5.34) | -0.28 (5.74) | 0.52 (5.57) | -0.93 (5.44) | 0.15 (5.21) | -0.75 (5.76) |
| SEVLT Recall change (mean (SD)) | 0.28 (2.95) | 0.28 (3.01) | -0.13 (2.89) | -0.10 (2.87) | -0.26 (2.93) | -0.01 (3.16) |
| Digits symbol change (mean (SD)) | -2.22 (7.41) | -2.22 (8.28) | -2.06 (6.93) | -2.51 (6.84) | -2.78 (7.93) | -0.26 (7.35) |

### PRS association with MCI

Based on each GWAS and genome-wide significant SNPs, we selected a single PRS by optimizing the CV across five independent subsets of the data. Figure 2 describes the association in terms of estimated odd ratios (OR) and 95% confidence intervals for MCI in the combined SOL-INCA dataset, and within restricted subsets of participants with at least 20% global European, African, or Amerindian genetic ancestry. Table 3 reports the clumping parameters and number of SNPs in the selected PRS according to each of the four GWAS. Surprisingly, selected PRS based on both FINNGEN and Jun et al. PRS were not clumped.

**Table 3:** Characteristics of the AD PRS selected in association with MCI in the SOL-INCA dataset.

| GWAS name | R<sup>2</sup> | Distance | # SNPs |
| --- | --- | --- | --- |
| FINNGEN | -- | -- | 168 |
| Jansen et al. | 0.3 | 500kb | 158 |
| Jun et al. | -- | -- | 203 |
| Kunkel et al. | 0.3 | 250kb | 122 |

For each GWAS, the table describes the characteristics of the PRS selected by optimizing the coefficient of variation across 5 independent subsets of the data. R<sup>2</sup> and “Distance” are parameters used for clumping. When they are not provided, the selected PRS used all available SNPs with p-value<5x10<sup>-8</sup>, without clumping.

**Figure 2:**
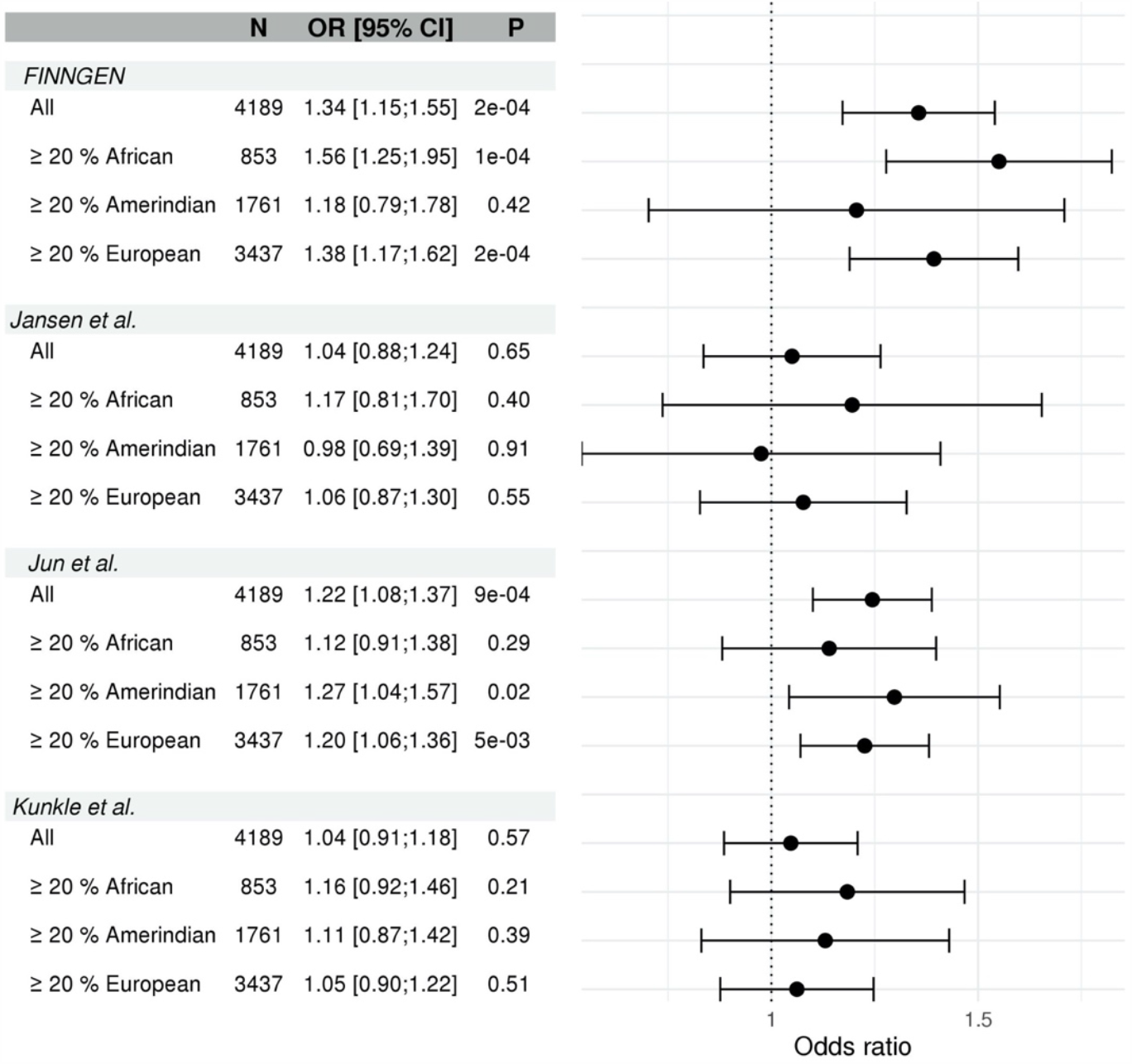
Estimated effect sizes and confidence intervals of PRS based on AD GWASs in association with MCI. For each GWAS, the PRS were selected based on optimizing the coefficient of variation across 5 independent subsets of the SOL-INCA dataset. We provide the effect size, confidence interval, and p-value in models based on the complete dataset (“All”), the subset of people with at least 20% global proportion of African, Amerindian, and European ancestries. The PRS association was estimated in a model adjusted for age at the HCHS/SOL baseline visit, time from HCHS/SOL baseline to the SOL-INCA visit, sex, study center, 5 principal components, and APOE-*ϵ*4 and APOE-*ϵ*2 allele counts.

As shown in Figure 2, two of the PRS were associated with MCI in the complete sample: FINNGEN PRS (OR=1.34, 95% CI [1.15, 1.55], p-value =0.0002), and PRS based on the multi-ethnic GWAS reported in Jun et al. (OR=1.22, 95% CI [1.08, 1.37], p-value = 0.0009). When removing 16 individuals with severe cognitive impairment/suspect dementia, the results were essentially the same (not shown). Supplementary Figure 1 describes the association of the PRS across a range of p-values, demonstrating that the estimated ORs are generally robust to the choice of p-value threshold, with the exception of FINNGEN PRS, in which the estimate OR becomes higher with higher p-value thresholds for SNPs inclusion. However, the CV also becomes higher for FINNGEN PRS with increasing CV, suggesting low reproducibility of associations across independent datasets.

Figure 2 further demonstrates that when restricting to individuals with at least 20% of a given ancestry, the estimated ORs with Jun et al. PRS remained about the same, while for FINNGEN, the estimated OR in the group of individuals with at least 20% global African ancestry was 1.56 (95% [1.25,1.95], p-value = 0.0001). Stratifying the association by Hispanic/Latino background group (Figure 3), shows that the FINNGEN PRS effect estimate was highest in the Cuban group (OR=1.68, 95% CI [1.31, 2.15]), and all group-specific effects were positive (risk increasing). However, the evidence of heterogeneity was weak (p-value = 0.22). In contrast, effect estimates from Jun et al. PRS in the Hispanic/Latino background groups showed higher heterogeneity (p-value = 0.03), with some of the effect estimates being negative. Specifically, the effect estimate for the South American group was protective (OR=0.6, 95% CI [0.39, 0.90], p-value=0.02). The highest estimated effect size was in the Puerto Rican group (OR=1.34, 95% CI [1.09, 1.66]). In secondary analysis, we also constructed a PRS as a weighted combination of FINNGEN and Jun et al. PRSs. The associations of this PRS with MCI in the combined dataset and in the Hispanic/Latino background groups are reported in Supplementary Figure 3. The OR was 1.44 (95% CI [1.25, 1.66], p-value = 4×10^−7^), and the largest estimated effect size was again in the Cuban background group (OR=1.71, 95% CI [1.33, 2.21]). There was no evidence for heterogeneity (p-value=0.49).

**Figure 3:**
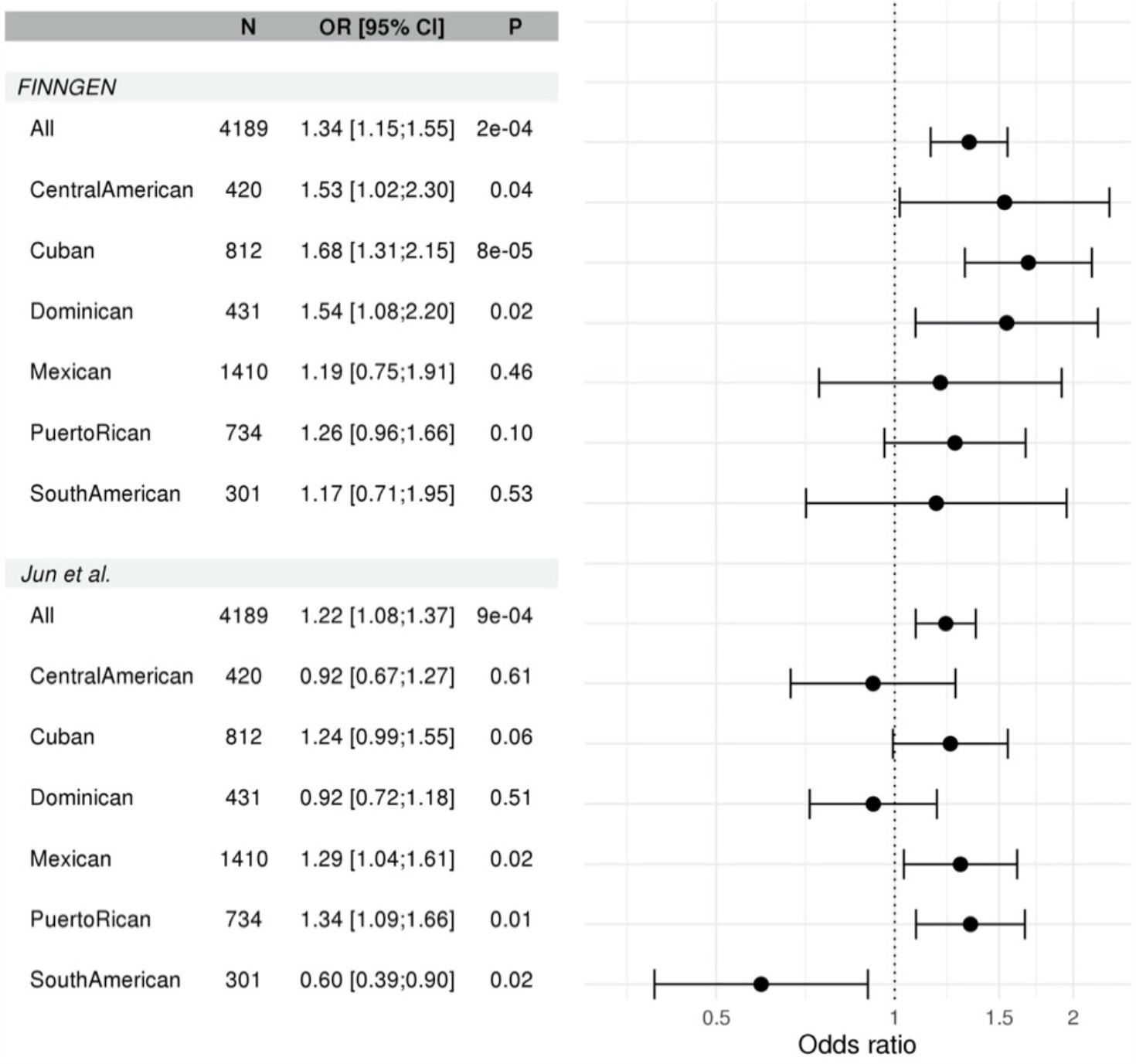
Association of MCI-associated AD PRS with MCI within Hispanic/Latino background group.

### PRS association with change in cognitive decline

Figure 4 provides effect estimates of global cognitive decline (G-factor change) and in change in specific cognitive tests. While all effect sizes were negative (pointing at decreased cognitive function for higher values of the AD PRS), none of the associations where statistically significant.

**Figure 4:**
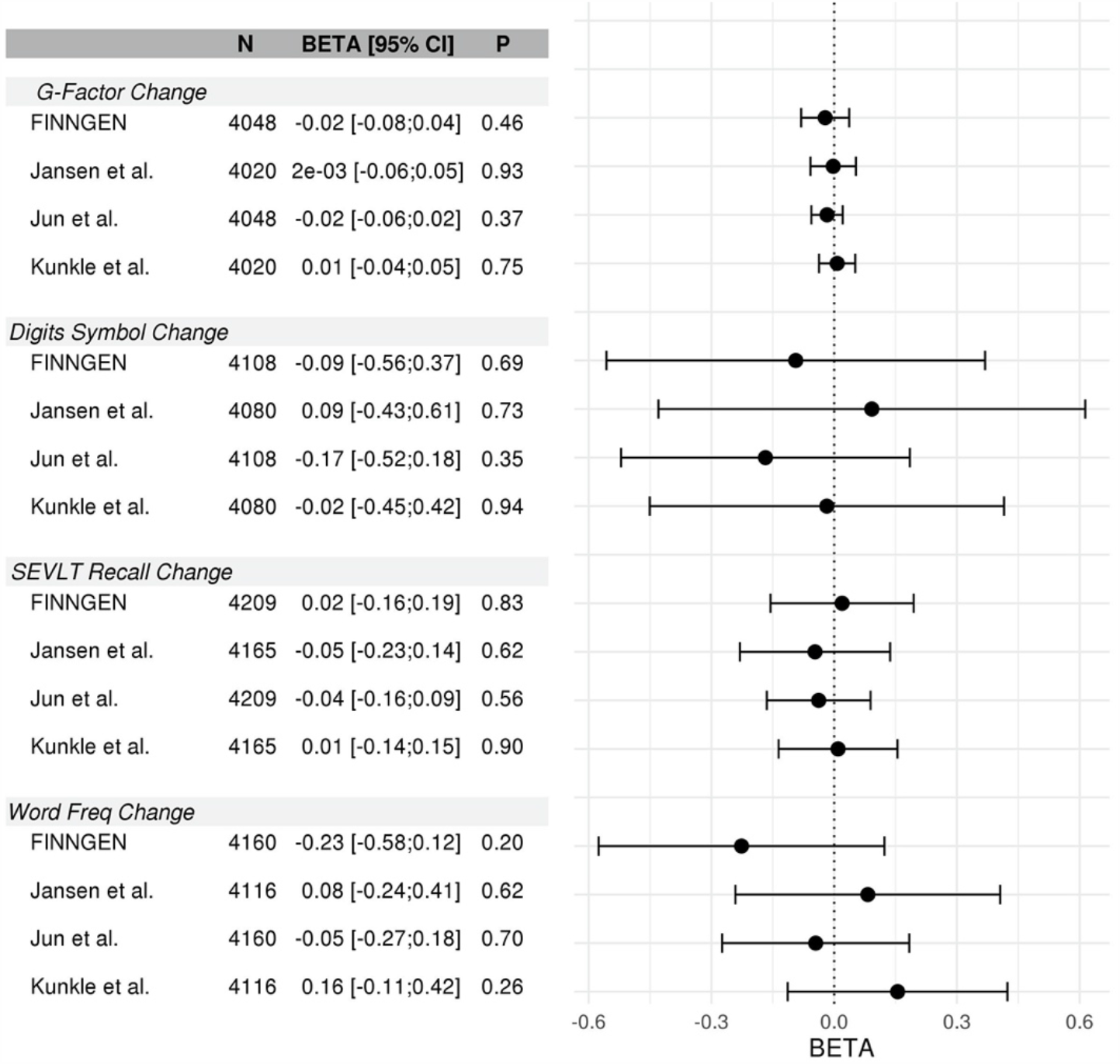
Association of the MCI-associated AD PRS with change in cognitive function.

### Relationship between APOE and AD PRS

Table 4 reports the association of the AD PRS and of *APOE*-*ϵ*4 and *APOE*-*ϵ*2 allele counts, in a model that accounted for all these genetic components together, with MCI. *APOE*-*ϵ*4 and *APOE*-*ϵ*2 alleles were generally unassociated with MCI, as we previously shown in this population (Granot-Hershkovitz et al. 2020). However, in the model with FINNGEN PRS, both *APOE* alleles were protective against MCI. When removing SNPs from 158 SNPs from the 1Mb region around the *APOE* rs429358 SNP, both *APOE* alleles associations and the PRS (now based on 10 SNPs, still all from chromosome 19) association were not statistically significant. This indicates co-linearity between *APOE* alleles and the PRS. Indeed, Table 4 also reports analysis of PRS associations without adjustment for *APOE* alleles. While FINNGEN PRS is still associated with increased risk of MCI, the effect is lower (OR=1.11, p-value=0.04). Supplementary Figure 2 further provides a forest plot of the estimated association of the FINNGEN PRS when removing the *APOE* region SNPs from the PRS, demonstrating the lack of association.

**Table 4:**
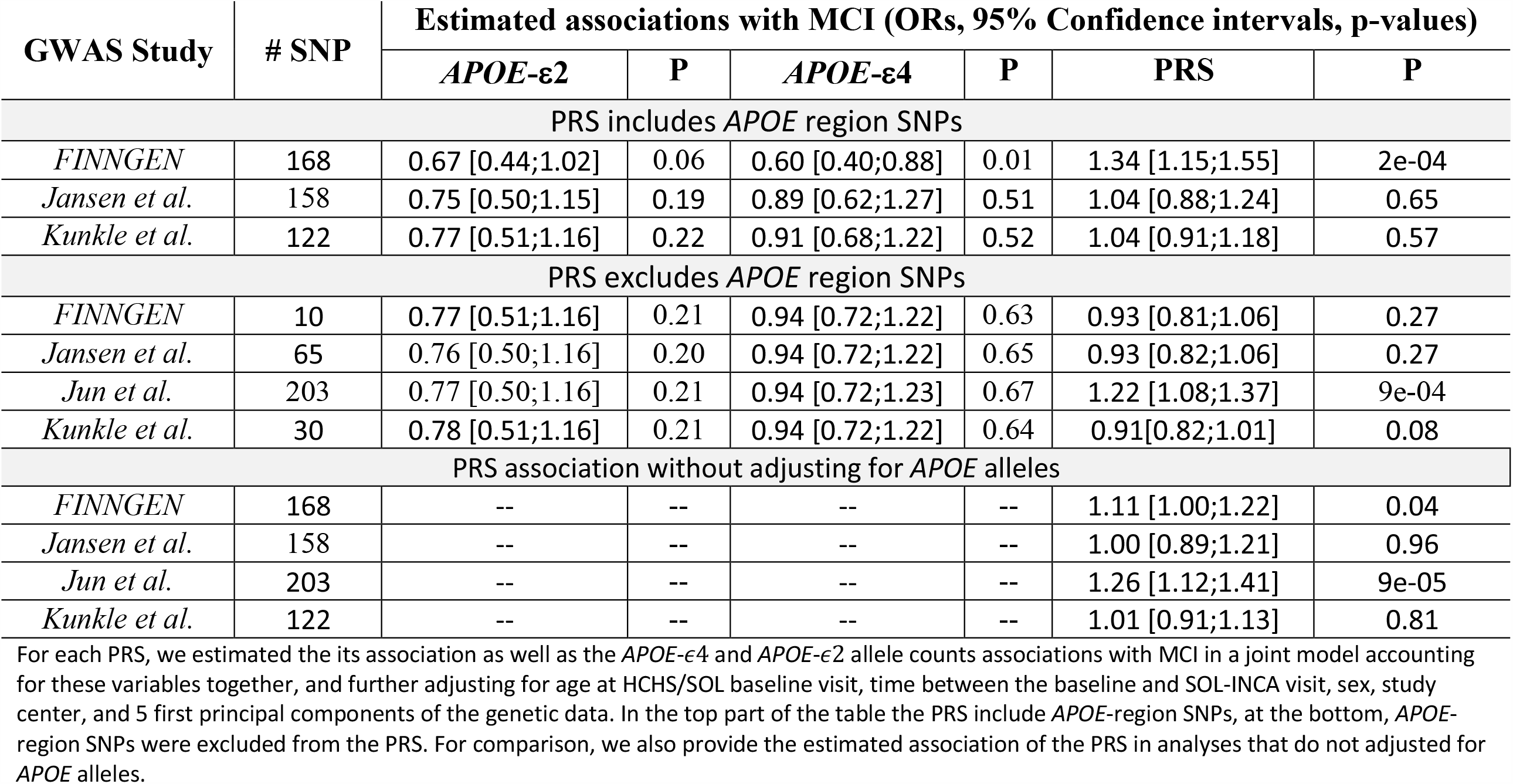
estimated association of AD PRS and of *APOE* allele counts in with MCI Estimated associations with MCI (ORs, 95% Confidence intervals, p-values)

Supplementary Figure 3 further studies the relationship of the FINNGEN based PRS with the *APOE*-*ϵ*4 allele, in the form of a PRS density within *APOE*-*ϵ*4 carriers and non-carriers. These densities are quite different, and within *APOE*-*ϵ*4 carriers the FINNGEN PRS has higher values, on average, compared to the PRS values in *APOE*-*ϵ*4 non-carriers.

Supplementary Table 1 reports the association of the primary AD PRS, without removing *APOE* region SNPs, with MCI within groups of *APOE*-*ϵ*4 carriers (n=1,358) and non-carriers (n=3,243). The ORs of the FINNGEN and Jun et al. PRS are similar in the two groups.

### Investigation of differences in MCI associations of different PRS

We compared the genomic regions represented by the four compared PRSs, as reported in Supplementary Figure 6. All genomic regions represented by the Jun et al. PRS and by FINNGEN PRS are also represented by the Kunkle et al. and Jansen et al. PRS. Both latter GWAS had additional genomic regions represented. We performed additional analyses to study why PRS based on both Kunkle et al. (2019) and Jansen et al. (2019) where not associated with MCI while PRS based on FINNGEN, another European-ancestry population, was associated.

Supplementary Table 2 report results from association analysis of PRS that used summary statistics from Kunkle et al. (2019) and Jansen et al. (2019), mimicking FINNGEN PRS by restricting SNP selection to SNPs from the two genomic regions on chromosome 19 represented in FINNGEN PRS. These chromosome 19-restricted PRS were associated with MCI in SOL-INCA (p-value=0.05 for both PRS), with lower OR compared to FINNGEN PRS. (OR=1.11, and 1.15 for Kunkle at al. PRS and Jansen et al. PRS, respectively, compared to OR=1.34 of FINNGEN PRS).

## Discussion

We studied the association between multiple PRS for AD and MCI in the SOL-INCA study of diverse U.S. Hispanics/Latinos. We used four GWAS to construct PRS: the latest published IGAP GWAS, a GWAS from FINNGEN, based on the Finland Biobank, a meta-analysis GWAS of AD and of AD-by-proxy, and a multi-ethnic GWAS. Both the FINNGEN-based PRS and the PRS based on the multi-ethnic GWAS in Jun et al. were associated with MCI in SOL-INCA. Interestingly, FINNGEN PRS was constructed almost entirely of SNPs in the *APOE* gene region, while the Jun et al. PRS did not have any SNPs in the *APOE* gene region. The two PRS were uncorrelated, and therefore, in a secondary analysis, we combined them into a single PRS using weights estimated in a logistic regression. Surprisingly, while the FINNGEN PRS was mostly comprised of *APOE* region SNPs and had different distribution among *APOE*-*ϵ*4 carriers and non-carriers, it was associated with MCI while the *APOE*-*ϵ*4 allele was not. None of the PRS were associated with cognitive decline phenotypes.

PRS based on both Kunkle et al. (2019) and Jansen et al. (2019) were not associated with MCI in the SOL-INCA. It is somewhat surprising that FINNGEN-based PRS was associated with MCI, given that it has lower sample size. We used both Kunkle et al. and Jansen et al. GWAS to construct PRS mimicking the FINNGEN PRS: we constructed PRS based only on SNPs from FINNGEN PRS association regions on chromosome 19, and including all genome-wide significant SNPs, with no clumping (Supplementary Table 2). These chromosome 19-restricted PRS had weaker association with MCI in SOL-INCA compared to FINNGEN PRS, suggesting that either the SNP selection or their effect sizes were less useful based on Kunkle et al. (2019) and Jansen et al. (2019). In comparisons of genomic regions represented by the various PRS, both Kunkle et al. (2019) and Jansen et al. (2019) GWAS PRS had additional genomic regions represented (compared to FINGENN and Jun et al.), possibly adding signal that does not strongly predict neurocognitive decline in middle-to later-aged Hispanics/Latinos. It is a topic of future work to further evaluate associations of specific genomics regions in Hispanics/Latinos throughout the aging process and to use this information to construct better PRS targeted to Hispanics/Latinos.

Two other studies specifically looked at the association of AD PRS with cognitive decline and MCI in middle-aged individuals, i.e. in similar age groups to the SOL-INCA cohort. Logue et al. (2019) considered AD PRS to predict MCI in non-Hispanic European ancestry individuals, and reported similar associations to those we observed: OR values between 1.17 to 1.4 (considering multiple p-value thresholds for including SNPs in the PRS) comparing cognitively normal adults and individuals with amnesic MCI. They also studied non-amnesic MCI, for which the association was weaker, suggesting heterogeneity of AD-related genetic association by type of MCI.

Like others, we also report associations for a range of p-value thresholds as a secondary analysis (Supplementary Figure 1). For FINNGEN- and Jun et al.-based PRS the association was strongest with the genome-wide significance threshold. Even for the genome-wide significance threshold there are choice to make regarding clumping parameters. To choose clumping parameters, we split the data to 5 non-intersecting, independent datasets, computed the effect size estimates in each, and computed a CV across these estimates. We selected the clumping parameters that optimized this CV. Thus, we believe that our estimated ORs are relatively robust against overfitting. Surprisingly, for both FINNGEN and Jun et al., the selected clumping parameters resulted in no clumping. This is surprising because many of the SNPs in the same genomic region are highly correlated. We think that the PRS without clumping were selected due to LD differences, as follows. Clumping means that the SNP with lowest p-value in the discovery GWAS, among many correlated SNPs, will be selected into the PRS. In a different population a different SNP may have a stronger association with the outcome, resulting in non-optimal SNP selection with clumping. Future work should identify a minimal set of SNPs that should be used from each distinct association region. It is plausible that larger data sets are required for achieving this goal without overfitting.

The PRS constructed by Logue et al. (2019), as well as by others, as reviewed in the introduction, were based on earlier IGAP GWAS (Lambert et al. 2013), from 2013, while we used an IGAP GWAS from 2019 (Kunkle et al. 2019), in addition to a few other GWASs. The PRS based on the IGAP GWAS were not associated with MCI nor cognitive decline in our data. While this could be due to differences in genetic ancestries, or to the different outcome used, the FINNGEN-based PRS was associated with MCI, despite also being based on GWAS in a European ancestry. Perhaps the greater homogeneity of the Finnish population and the consistency in phenotype definition, compared to the IGAP meta-analysis, increased the precision of the effect size estimates in FINNGEN compared to the IGAP GWAS that we used, resulting in a more powerful PRS.

An important question is whether our findings explain, in part, disparities in AD and MCI in Hispanics/Latinos, compared to European ancestry individuals and within Hispanics/Latinos of diverse backgrounds. While we still cannot answer this question, an important observation is that the FINNGEN PRS, composed of mostly SNPs in the *APOE* gene region, has stronger effect in individuals with high proportion of African ancestry. At the same time, the *APOE*-*ϵ*4 allele count was not associated with MCI while the FINNGEN PRS was. This adds to a cumulative evidence about differences in the *APOE* gene region architecture and association with cognitive aging phenotypes by genetic ancestry (Blue et al. 2019, Cornejo, Granot-Hershkovitz et al. 2020, Teruel et al. 2011). Other investigations focused on the *APOE* alleles themselves, this is the first time where we see evidence of heterogeneity in a combination of *APOE*-related SNPs, rather than *APOE*-*ϵ*4 and *ϵ*2 alleles. Disparities in AD in Hispanics/Latinos are likely, at least in part, due to disparities in environmental and sociological exposures, such as air pollution (Kulick et al. 2020), or socioeconomic status (Sheffield & Peek 2009), which is potentially associate with many environmental and psychological factors. These disparities may be associated with differences in genetic risk, and in gene-environment interaction, where environmental exposure exacerbate genetic risk. In future research we will use these PRS to study how environmental exposures modify the PRS effect on MCI and AD.

Specific strengths of our study are the use of well phenotyped, yet understudied, diverse Hispanics/Latinos, comprehensive genetic data including proportions of global ancestries, and the use of multiple GWAS to construct PRS. Our study also has a few limitations. First, the MCI trait was not based on biological biomarkers, but rather on the NIA-AA criteria. Among people with cognitive performance from 1 to 2 SD below the mean of their peers, there may be inclusion of individuals with life-long below average cognitive performance who are not on a trajectory of cognitive decline. In longitudinal studies, many individuals who meet a clinical case definition for MCI one year revert to normal the next (Thomas et al. 2019). In addition, not all MCI is attributable to Alzheimer’s disease. Some individuals may have MCI attributable to vascular disease or other pathologic substrates. If the MCI case group includes many individuals who do not have MCI attributable to Alzheimer’s disease, that could lead to underestimation of the relative odds of MCI given a PRS and attenuate our ability to optimally identify PRS. Second, cognitive trajectories were estimated based on two points in time using cognitive tests with modest retest reliabilities. The lack of association of PRS with indices of cognitive decline may reflect unreliability in the estimated slopes. An additional wave of follow-up data may strengthen our estimates of slope and our ability to identify PRS linked to cognitive trajectory. Third, while we were able to investigate the heterogeneity of the PRS across Hispanic/Latino background and genetic ancestry groups, we could not form a specific recommendation regarding optimal PRS by background or ancestral characteristics of an individual, due to small sample sizes. Future work will ideally study the specific SNPs and their effects across genetic ancestries, to obtain more precise PRS adapted to individuals by their ancestries, and also to pin-point the specific SNPs that may have different frequencies and perhaps different effects across ancestries.

In summary, we used summary statistics from AD GWAS to construct multiple PRS, two of which were associated with MCI in U.S. Hispanics/Latinos. Surprisingly, none of the PRS was associated with cognitive decline. In contrast, the *APOE*-*ϵ*4 allele was associated with cognitive decline but was not associated with MCI in SOL-INCA (Granot-Hershkovitz et al. 2020). As SOL-INCA participants age, it will be important to study whether this pattern continues or whether the observed cognitive decline will become more strongly related to AD development. Cognitive decline may be the results of other health and disease phenotypes, such as diabetes and kidney function. PRS could be developed for such phenotypes, and also better adapted to Hispanics/Latinos, due to larger availability of GWAS in diverse populations. In future work we will study genetic prediction of cognitive decline in Hispanics/Latinos using PRS for risk factors for cognitive decline, and PRS associations while accounting for lifestyle and other risk factors.

## Data Availability

Genotype and phenotype data from the Hispanic Community Health Study/Study of Latinos can be obtained by application to dbGaP, accession phs000810.v1.p1.

## Acknowledgements

The authors thank the staff and participants of HCHS/SOL for their important contributions. Investigators website - http://www.cscc.unc.edu/hchs/.

