## Supplementary materials for "Polygenic Risk Scores for Alzheimer’s Disease and Mild Cognitive Impairment in Hispanics/Latinos in the U.S: The Study of Latinos – Investigation of Neurocognitive Aging"

Sofer et al.

**Supplementary Figure 1: MCI associations of PRS based on varying p-value thresholds.**

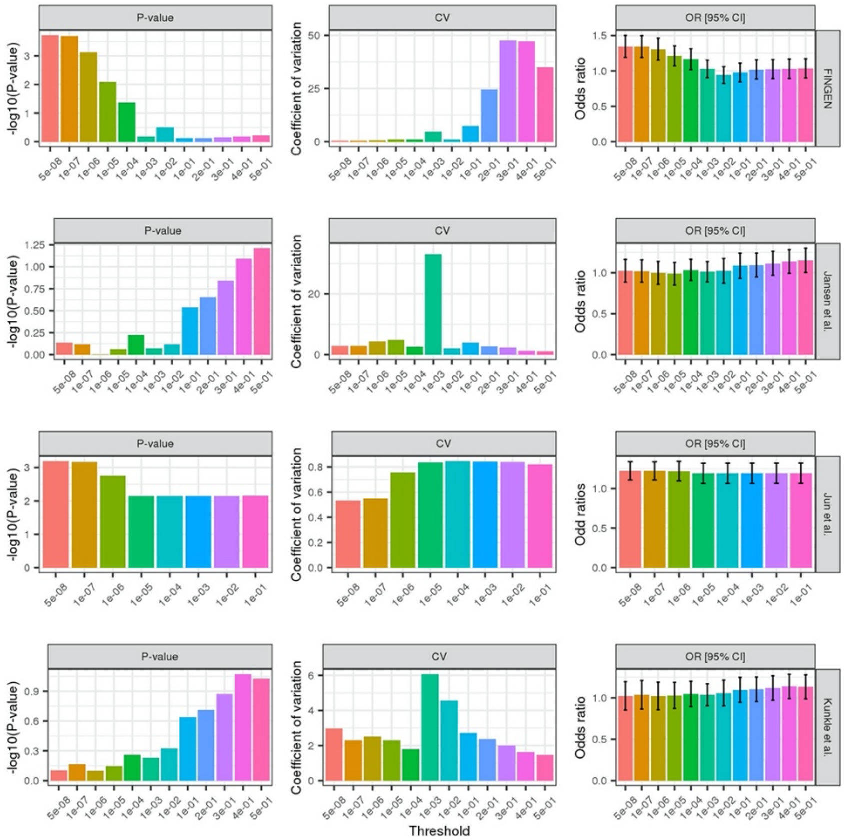


For each of the 4 GWAS described in Table 1 in the main manuscript, we constructed PRS based on a range of p-value threshold, using the clumping parameters selected in the primary analysis. Performance of each PRS are described: -log10(p-value) of the association with MCI in the combined SOL-INCA cohort, coefficient of variation (CV) computed based on effect estimates in 5 independent subsets of the data, and estimated odds ratios (OR) and 95% confidence intervals.

**Supplementary Figure 2: Estimated effect sizes and confidence intervals of FINNGEN AD PRS after removing SNPs from the *APOE* gene region**

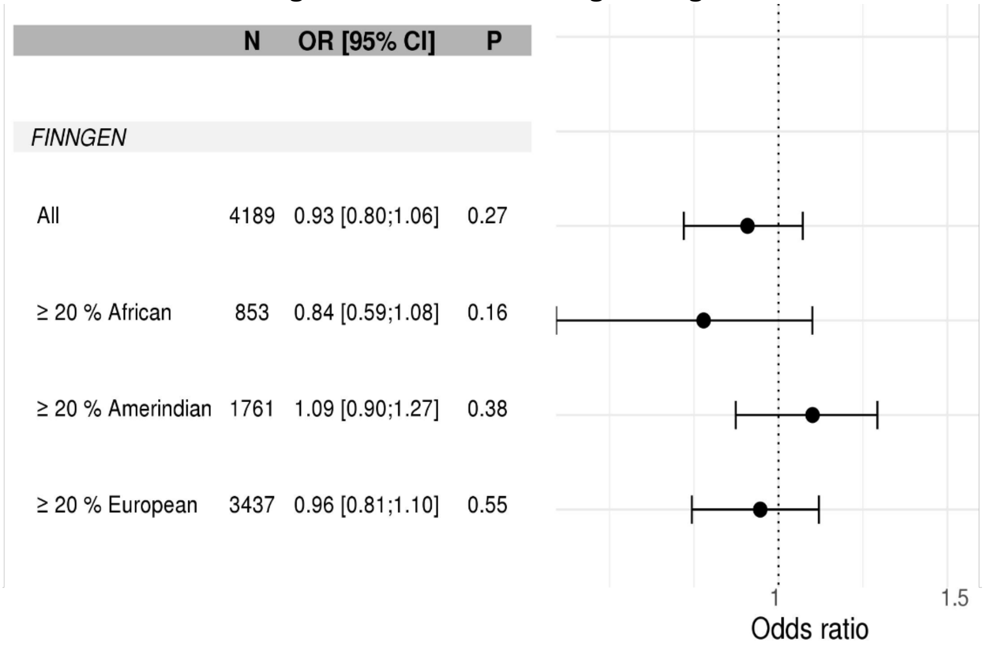


For the same PRS reported in the main analysis (Figure 1), we excluded SNPs from the *APOE* gene region (SNPs 1000Kb around the SNP rs429358). We provide the effect size, confidence interval, and p-value in models based on the complete dataset ("All"), the subset of people with at least 20% global proportion of African, Amerindian, and European ancestries. The PRS association was estimated in a model adjusted for age at the HCHS/SOL baseline visit, time from HCHS/SOL baseline to the SOL-INCA visit, sex, study center, 5 principal components, and *APOE*-$\epsilon4$ and *APOE*-$\epsilon2$ allele counts. The PRS based on Jun et al.'s GWAS did not include *APOE* region SNPs and is presented here only for comparison with FINNGEN PRS.

**Supplementary Figure 3: Estimated effect sizes and confidence intervals of AD PRS constructed based on a joint model of FINNGEN PRS and Jun et al. PRS**

| | N | OR [95% CI] | P |
|---|---|---|---|
| *FINNGEN + Jun et al.* | | | |
| All | 4195 | 1.44 [1.25;1.66] | 4e-07 |
| CentralAmerican | 420 | 1.17 [0.83;1.65] | 0.37 |
| Cuban | 815 | 1.71 [1.33;2.21] | 6e-05 |
| Dominican | 431 | 1.36 [0.94;1.96] | 0.11 |
| Mexican | 1412 | 1.47 [1.00;2.16] | 0.05 |
| PuertoRican | 734 | 1.49 [1.16;1.90] | 2e-03 |
| SouthAmerican | 301 | 0.71 [0.41;1.21] | 0.21 |

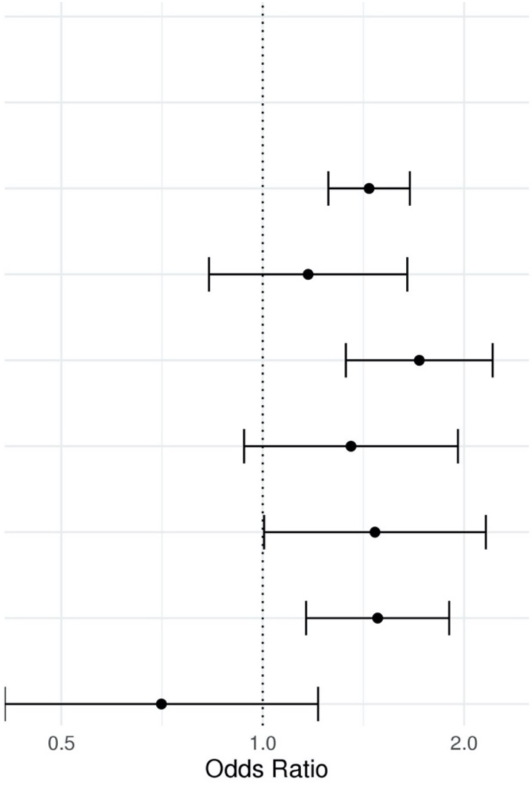


The PRS was constructed as a weighted combination of FINNGEN PRS and Jun et al. PRS. We provide the effect size, confidence interval, and p-value in models based on the complete dataset ("All"), and by Hispanic/Latino background groups. The PRS associations were estimated in a model adjusted for age at the HCHS/SOL baseline visit, time from HCHS/SOL baseline to the SOL-INCA visit, sex, study center, 5 principal components, and *APOE*-ϵ4 and *APOE*-ϵ2 allele counts.

**Supplementary Figure 4: Distribution of FINNGEN PRS within *APOE*-$\epsilon 4$ carriers and non-carriers.**

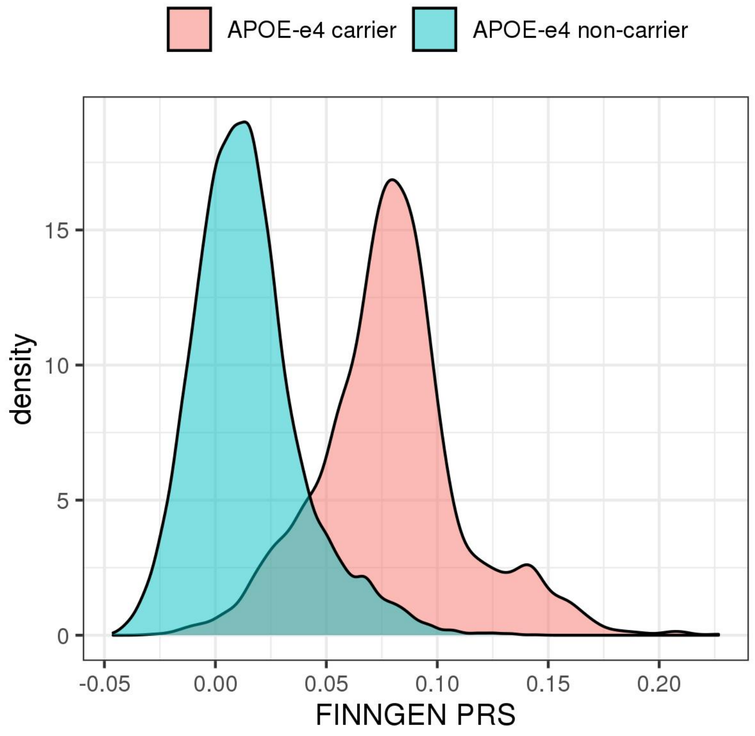

**Supplementary Figure 5: Estimated effect sizes and confidence intervals of PRS based on AD GWASs in association with MCI, excluding 16 individuals with severe impairment/suspect dementia**

| | N | OR [95% CI] | P |
|---|---|---|---|
| *FINNGEN* | | | |
| All | 4177 | 1.35 [1.16;1.57] | 1e-04 |
| ≥ 20 % African | 852 | 1.57 [1.26;1.96] | 9e-05 |
| ≥ 20 % Amerindian | 1759 | 1.19 [0.79;1.79] | 0.42 |
| ≥ 20 % European | 3428 | 1.38 [1.17;1.63] | 2e-04 |
| *Jansen et al.* | | | |
| All | 4177 | 1.02 [0.86;1.22] | 0.80 |
| ≥ 20 % African | 852 | 1.17 [0.81;1.70] | 0.41 |
| ≥ 20 % Amerindian | 1759 | 0.98 [0.69;1.40] | 0.93 |
| ≥ 20 % European | 3428 | 1.04 [0.85;1.28] | 0.71 |
| *Jun et al.* | | | |
| All | 4177 | 1.24 [1.10;1.39] | 3e-04 |
| ≥ 20 % African | 852 | 1.13 [0.91;1.39] | 0.27 |
| ≥ 20 % Amerindian | 1759 | 1.27 [1.03;1.57] | 0.02 |
| ≥ 20 % European | 3428 | 1.22 [1.07;1.38] | 3e-03 |
| *Kunkle et al.* | | | |
| All | 4177 | 1.04 [0.91;1.19] | 0.57 |
| ≥ 20 % African | 852 | 1.18 [0.91;1.54] | 0.22 |
| ≥ 20 % Amerindian | 1759 | 1.13 [0.89;1.42] | 0.32 |
| ≥ 20 % European | 3428 | 1.06 [0.91;1.24] | 0.42 |

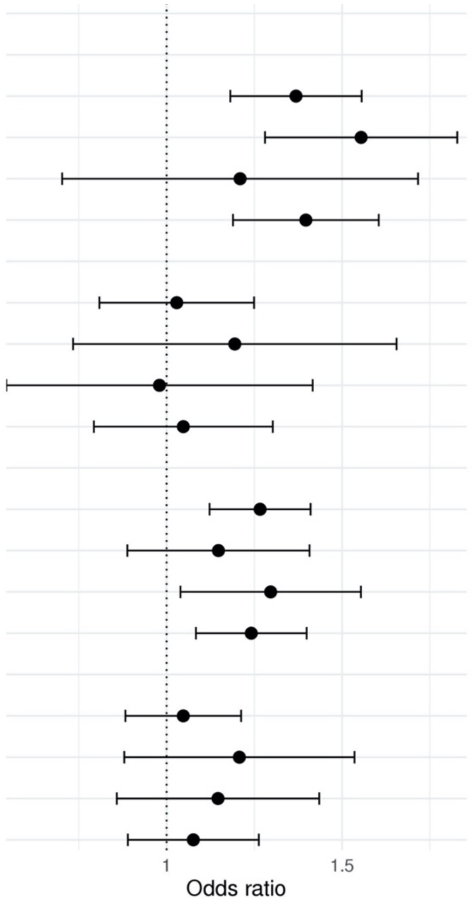


The association reported here mimics exactly those in reported in Figure 1 of the main manuscript, with the exception excluding the 16 individuals with sever impairment/suspect dementia.

**Supplementary Figure 6: Venn Diagram demonstrating the overlap in genomic regions represented by each of the compared PRS.**

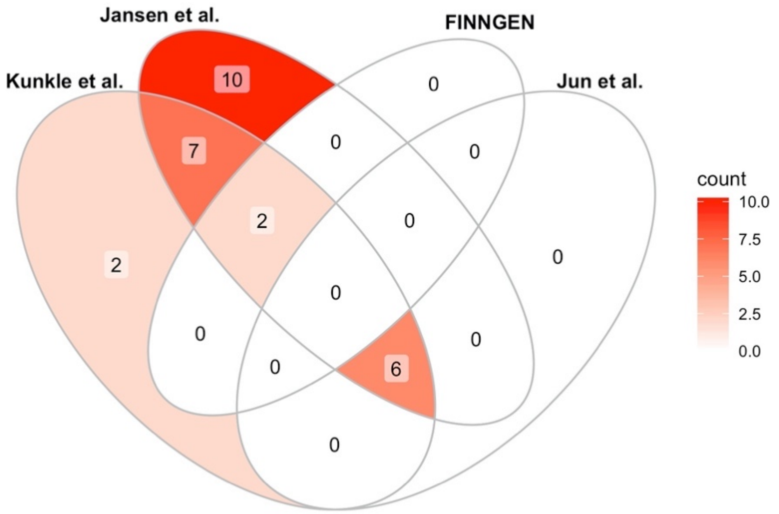

**Supplementary Table 1: PRS association with MCI within APOE-$\epsilon 4$ carriers (n=1,358) and non-carriers (n=3,243).**

| ***GWAS Study*** | **# SNP** | ***APOE-$\epsilon 4$* carriers** | | ***APOE-$\epsilon 4$* non-carriers** | |
|---|---|---|---|---|---|
| | | **OR (95% CI)** | **P** | **OR (95% CI)** | **P** |
| *FINNGEN* | 168 | 1.25 [1.01;1.54] | 0.04 | 1.19 [1.06;1.33] | 3e-03 |
| *Jansen et al.* | 89 | 1.07[0.88;1.32] | 0.50 | 1.03 [0.89;1.20] | 0.68 |
| *Jun et al.* | 203 | 1.25 [1.00;1.56] | 0.05 | 1.26 [1.12;1.43] | 2e-04 |
| *Kunkle et al.* | 235 | 0.96 [0.79;1.18] | 0.72 | 1.08 [0.93;1.25] | 0.31 |

Association of the PRS reported in Figure 2 when stratifying by *APOE*-$\epsilon 4$ carrier status. The PRS associations were estimated in a model adjusted for age at the HCHS/SOL baseline visit, time from HCHS/SOL baseline to the SOL-INCA visit, sex, study center, and 5 principal components (and not adjusting for *APOE*-$\epsilon 4$ and *APOE*-$\epsilon 2$ allele counts).

**Supplementary Table 2: Associations of chromosome 19 restricted PRS based on Kunkle et al. and Jansen et al. GWASs with MCI.**

| GWAS Study | # SNP | Estimated associations with MCI (ORs, 95% Confidence intervals, p-values) | | | | | |
|---|---|---|---|---|---|---|---|
| | | ***APOE*-ε2** | **P** | ***APOE*-ε4** | **P** | **PRS** | **P** |
| PRS includes *APOE* region SNPs | | | | | | | |
| *Jansen et al.* | 610 | 0.73 [0.49;1.10] | 0.13 | 0.77 [0.56;1.10] | 0.11 | 1.16 [1.00;1.34] | 0.05 |
| *Kunkle et al* | 499 | 0.76 [0.51;1.15] | 0.20 | 0.85 [0.64;1.13] | 0.26 | 1.12 [1.00;1.26] | 0.05 |

The PRS were constructed with no clumping, based on genome-wide significant SNPs on chromosome 19, mimicking the FINNGEN PRS. The analyses were adjusted for age at the HCHS/SOL baseline visit, time from HCHS/SOL baseline to the SOL-INCA visit, sex, study center, 5 principal components, and *APOE*-$\epsilon 4$ and *APOE*-$\epsilon 2$ allele counts.
